# High-resolution computational modeling of Transcranial Photobiomodulation: Light propagation and thermal effects

**DOI:** 10.1101/2025.01.18.25320758

**Authors:** Alexander R Guillen, Dennis Q Truong, Paula Cristina Faria, Brian Pryor, Luis de Taboada, Abhishek Datta

## Abstract

**Background:** Transcranial Photobiomodulation (tPBM) is the non-invasive application of light to modulate underlying brain activity. There is increasing interest in evaluating tPBM as a therapeutic option. The typical technological questions are extent of light penetration and associated tissue temperature increases. Limited computational efforts to quantify these aspects are restricted to simplified models.

**Methods:** We consider a 3D high-resolution (1 mm) anatomically realistic head model to simulate tPBM at 800 nm wavelength and using power densities spanning three decades (10, 100, and 1,000) mW/cm^2^. The intended target was the F3 region. We also tested time-variant application at 100 mW/cm^2^ for up to 20 minutes. Finally, tissue temperature rises for the American National Standards Institute (ANSI) safety limit of 330 mW/cm^2^ was also investigated at a test case.

**Results:** Our predictions reveal that the induced cortical irradiance is largely focal demarcated by the shape and extent of the source. Around 1% of the injected irradiance reaches the gray matter. Aligned with previous efforts, the scalp accounts for the greatest loss (∼65%). The irradiance reduces to a hundredth of the value from gray matter at ∼113 mm perpendicular distance from its surface. There is a growing halo-like effect at the level of CSF which is extended down to the underlying cortex. The CSF was found to be mainly responsible for this effect. We observe scalp temperature increases of 0.38°C and 3.76°C for 100 and 1,000 mW/cm^2^ power density, respectively. The corresponding brain temperature increases are predicted to be 0.06°C and 0.57°C. As expected, irradiance absorption is linear with applied power density. While maximum induced scalp temperature increases linearly with power density, maximum brain temperature increases less slowly with power density. Transient analysis at 100 mW/cm^2^ power density demonstrates expected scalp temperature rise with increasing stimulation duration. Temperature rises asymptote in ∼ 10 minutes.

**Conclusions:** tPBM presents unique potential to directly impose desired spatial profile using simple alteration of shape and size of the source. Usage of power density of 1,000 mW/cm^2^ exceeds scalp and brain temperature safety limits. Contrary to prior reports, light penetration can exceed >10 cm from gray matter surface.

## 1. Introduction

Transcranial photobiomodulation (tPBM) is an emerging non-invasive therapy involving the application of low-intensity light in the red to near-infrared (NIR) spectrum to stimulate neural tissue. First discovered by Mester et al. in 1968^1^, tPBM operates within wavelengths of 600 to 1,100 nm and energy fluences typically ranging from 1 to 20 J/cm². The mechanism of action is hypothesized to involve upregulation of cytochrome c oxidase (CCO) activity in mitochondria, promoting ATP production and enhancing cerebral oxygenation^2,3^. Despite promising preliminary results in neurodegenerative disorders^4,5^, traumatic brain injury^6^, and mood regulation^7^, optimal parameters for clinical efficacy and safety remain undefined. This study aims to bridge existing knowledge gaps by leveraging advanced finite element method (FEM) modeling to simulate light distribution and thermal effects in a high-resolution 3D anatomical model. We specifically evaluate light propagation, absorption profiles, and temperature increases under varying power densities, providing insight into safe and effective dosing for tPBM.

tPBM is currently in its nascent stages of clinical application. To our knowledge, there are currently no US FDA or Europe (medical CE) approved devices based on tPBM. As far as safety is concerned, large trials involving vulnerable populations (e.g. encompassing >500 acute stroke patients) have reported no significant disparity in side effects between active and sham stimulation^8^. In this study, an 808 nm laser was applied at 20 predetermined scalp locations, with each site receiving 2 minutes of stimulation, resulting in a 40-minute session with an energy dose of 1 J/cm^2^ per location. An extended duration investigation focused on Major Depression (20-30 minutes per session, twice weekly over 8 weeks) noted mild and transient side effects including headaches, insomnia, irritability, and unusual taste sensations^9,10^. The trial delivered NIR light at 823 nm as a continuous wave to the patients’ F3 and F4 EEG sites with a power density of 36.2 mW/cm^2^, equating to an energy dose of up to 65.2 J/cm^2^ per session. While employing different doses, tPBM applications across other clinical populations (TBI, Autism Spectrum Disorder, Dementia) have reported mild transient to none adverse events ^6,11–13^

The exact power density threshold required to achieve beneficial biological effects in the brain remains an active area of research^14^. Recommendations vary widely, adding complexity to the understanding of optimal parameters. Some groups advocate for power densities below 100 mW/cm^2^, with energy densities ranging from 4 to 10 J/cm^2^ at the target tissue level ^15^. Conversely, others propose higher energy densities, reaching up to 50 J/cm^2^ at the tissue surface^11^. In light of this, it is important to note that the application of PBM leverages two key metrics, *power density (irradiance)* and *energy density (fluence)*. Although both metrics measure energy intensity per unit area, they differ in application. Power density, the amount of power delivered per unit area, is particularly significant in continuous-wave (CW) laser applications. When high irradiance values above 1000 mW/cm^2^ are applied, it is standard practice to move the spot continuously to prevent tissue overheating^16^. Zein et al., 2018^15^ report that exceeding the following irradiance levels may cause tissue heating and potential photothermal effects: ∼ 100 mW/cm^2^ at 400-500 nm, ∼ 300 mW/cm^2^ at 600-700 nm, and ∼ 750 mW/cm^2^ at 800-900 nm. Fluence, incorporating duration, is critical in pulsed applications where timing must be precise to avoid excessive thermal build up that could compromise cellular integrity. Studies suggest that for PBM, fluences can vary between 0.03 and 100 J/cm^2^ at the target tissue^17^.

Given a transcranial technique, the extent of light penetration through the head is naturally an important dosing consideration. Several studies have sought to estimate experimentally the transcranial transmission of NIR and red light. In a study using human cadaver heads that included the skull with intact soft tissue, Jagdeo et al.^18^ found that the penetration of 830 nm light varied across anatomical regions of the skull (0.9% at the temporal region, 2.1% at the frontal region, and 11.7% at the occipital region). Notably, red light (633 nm) exhibited minimal penetration through the skull. Using human cadaver heads that also included scalp, Tedford et al, 2015^19^ found that 808 nm light could penetrate to a depth of 40-50 mm in the brain, compared to 660 nm and 940 nm light. Furthermore, Lapchak and colleagues reported NIR light transmission rates of ∼4% at 800 nm when applied to human cadaveric skulls ^20^. Recently, Morse et al.^21^ measured transmission of 750 and 940 nm light through 4 cm of fresh (unfixed) human cadaver heads, focusing on the frontal, parietal, occipital, and temporal lobes. Using a 4 W input power for each wavelength, they observed that the parietal lobe consistently exhibited the highest detected power across all four cadavers. The measured values in this region ranged from 77.2 to 358.1 µW for 750 nm and 134.4 to 395.2 µW for 940 nm^21^.

Computational studies have explored light penetration through the human head’s tissue layers. Stochastic based methods such as Monte Carlo modeling have been extensively used in earlier investigations^22–25^. The more recent approaches have employed FEM based approaches due its robustness and efficiency-especially when applied to complex geometries. While the geometries considered in the 3D FEM approaches have advanced over the recent years, they include simplifications. These range from fusing tissue compartments^26^, omitting CSF, with coarse gyri/sulci detail^27^. We perform here an extensive FEM guided analysis of light propagation and associated thermal effects using a high resolution highly accurate anatomical model. Using a single light source targeting the frontal cortex, we simulate tPBM across three power densities (10, 100, 1000 mW/cm^2^) and develop high fidelity visualization maps. We further explore time-variant application and influence of tissue properties. An in-depth analysis such as this, is expected to help researchers investigating this technique in many aspects-from improving understanding of the modality, shedding new insight, and to guide the rational dosing of future protocols.

Results from a preliminary investigation from this investigation were previously presented in a conference proceeding^28^.

## 2. Methods

We developed a high resolution 3D finite element method (FEM) model based on an average head MRI dataset (MNI152). tPBM was simulated by application of power densities spanning three decades targeting the F3 region. Light propagation simulation was based on the radiative transfer equation while thermal effects simulation based on the Pennes bio-heat equation. We considered both steady-state and time-variant application.

The exact simulation steps are outlined as follows:

### 2.1 Anatomical Dataset

The Caucasian brain atlas (ICBM-152) was obtained from the Montreal Neurological Institute (MNI, Montreal, Canada)^29^. It consists of 1 x 1 x 1 mm^3^ voxels of 256 gray levels. A representative average template was considered appropriate given the goal to shed light on the impact of tPBM that should hold across a large population. The impact of individual effects was not the focus of this study.

### 2.2 Image Processing and Segmentation

Segmentation of the MRI data into tissue categories was based on extensive prior study by our group ^30–32^. Briefly, the NIfTI (.nii) scan from the ICBM-152 dataset was processed using a combination of a probabilistic segmentation routine, tissue probability map, and a Matlab script to correct for segmentation errors. The resulting masks were imported into Simpleware (Synopsys Ltd., CA, USA) to correct anatomical and continuity errors using automated and manual tools.

### 2.3 Laser Placement and Meshing

Following tPBM administration, a cylinder with a radius of 20 mm and 10 mm in length (thickness) was placed at the F3 location on the scalp representing an applied beam area of 12 cm^2^ as illustrated in **Fig. 1**. The mesh was generated using an automatic free tetrahedral meshing process (+FE Free) with a fine coarseness setting of −16, tailored for finite element analysis in COMSOL Multiphysics 5.6 (COMSOL Inc., MA, USA).

**Figure 1:**
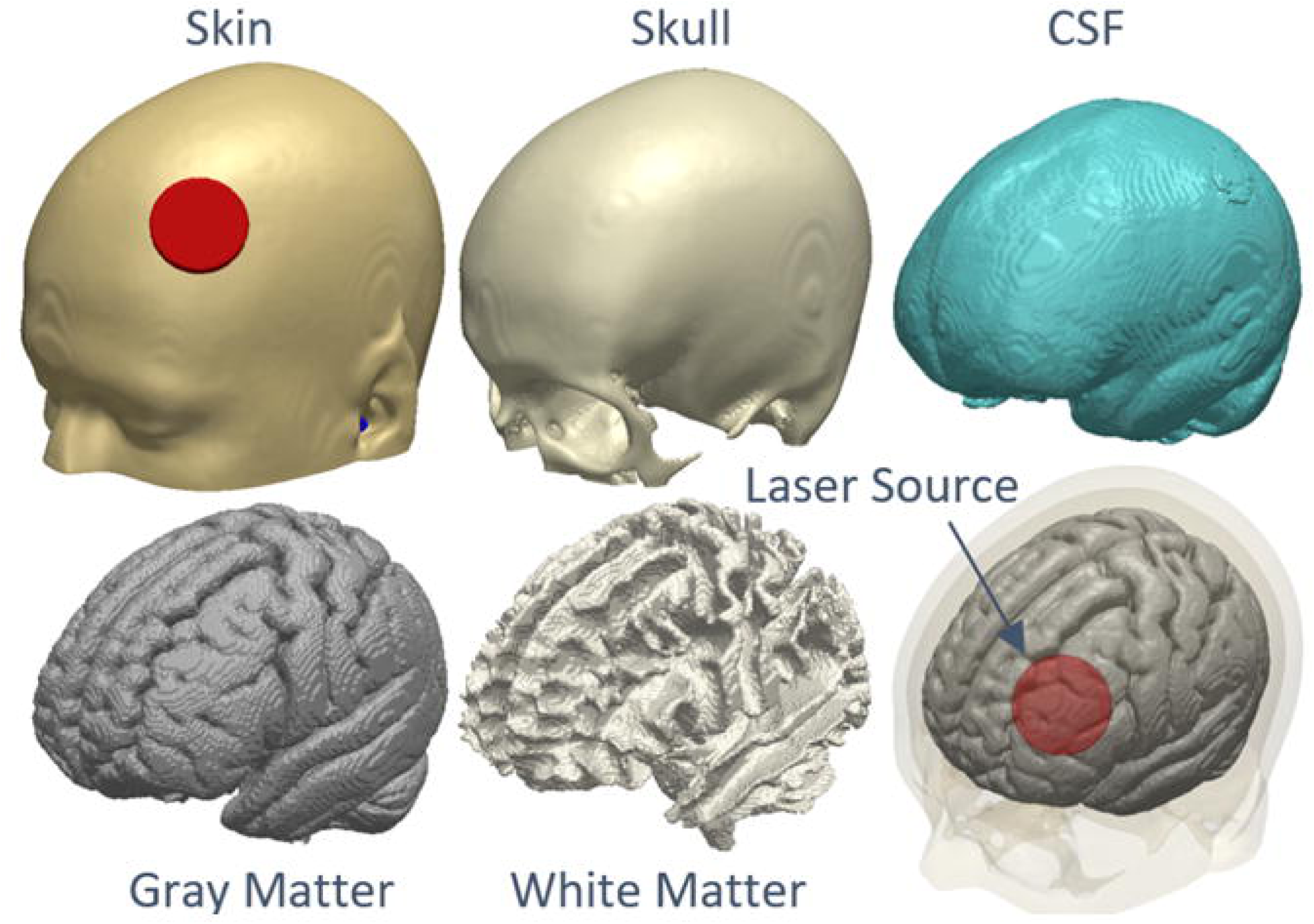
Segmentation of the MNI 152 template into tissue categories and laser source placement. The underlying cortical surface through the laser source is shown in the second row.

### 2.4 Finite Element Model

The volumetric mesh was imported into COMSOL to simulate tPBM administration. The mesh used in this study consisted of 3,406,015 vertices and 1,887,648 tetrahedral elements, supplemented by 482,840 triangular surface elements and 23,172 edge elements. The overall mesh volume was 3,745,000 mm^3^. The quality of the elements was assessed using the skewness metric, with values ranging from 0.05753 (minimum element quality) to 0.6088 (average element quality). The skewness measure ensures that elements are shaped in a way that minimizes numerical inaccuracies during simulations.

Within COMSOL, the simulation was configured for a 3D space dimension, utilizing multiphysics involving the Helmholtz equation (hzeq) and bioheat transfer (ht), under both stationary (CW laser application) and time dependent (PW laser application) study types.

#### 2.4.1 Light Propagation: Optical Properties and Model Configuration

A common framework for describing light diffusion in absorbing and scattering media is the radiative transfer equation (RTE), expressed as:

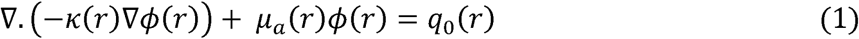

where (r) is the photon fluence rate (mW/cm^2^) at a radial distance r from the source, representing the power per unit area distributed within the medium, q_0_ (r) is the isotropic source distribution (mW/cm2) at r, and *K(r)* is the diffusion coefficient [3 (µ_α_ + µ′_s_)l-1 in centimeters (cm). Here, µ′_s_ (1/cm1) is the reduced scattering coefficient and µ_α_ (1/cm^1^) is the absorption coefficient.

To solve Equation (1), appropriate boundary conditions must be specified. At the interface between light source and tissue, the photon diffusion was constrained by the surface Dirichlet boundary condition, given by:

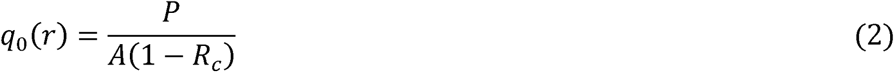

where P is the source power in watts (W), A is the beam area measured in square centimeters (cm^2^), and R_c_ is the epidermis reflection coefficient due to the refractive index (n) mismatch between air and skin. Thus, *1-R_c_* represents the fraction of incident energy that is absorbed by the tissue. The reflection coefficient for normal incidence can be computed using Fresnel’s equation:

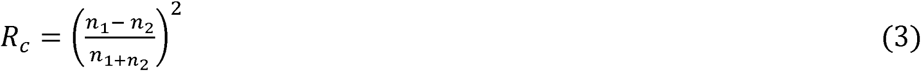

where n_1_ is the refractive index of the incident medium (e.g., air, typically n_1_ = 1) and n_2_ is the refractive index of the second medium (e.g., skin, typically n_2_ = 1.37)

For this study, the applied laser irradiance was 100 mW/cm^2^ for a source power of 1.79 W. Additional computations were performed at 10 mW/cm^2^ (0.18 W) and 1000 mW/cm^2^ (17.89 W).

At tissue-tissue interfaces, a Robin-type (flux/source) boundary condition was applied to maintain continuity of radiant energy in the normal direction. This condition guarantees a constant flux of energy perpendicular to the interface, irrespective of absorption, scattering, and refractive index variations between tissues. For CSF, boundary effects due to absorption and impedance were excluded because of its weak scattering properties ^33^. Weak scattering occurs when the refractive index mismatch between the scattering volume and the surrounding medium is negligible, reducing the strength of scattering events and allowing more light to pass through without significant deviation. Conversely, other tissues were modeled to minimize energy flux losses.

**Table I** presents the relevant optical properties of the modeled tissues^34,35^. These parameters depend on both spectral range (e.g., 800-880 nm) and skin pigmentation^36,37^. We assumed light skin pigmentation in the study. In addition, the tissue thickness (δ_tissue_) along the laser path has been incorporated to provide a comprehensive parameter list of the layered biological media.

**TABLE I.** OPTICAL PROPERTIES OF HEAD TISSUES (800-nm wavelength) AND TISSUE THICKNESS.

| Tissue | $\mu_a(1/\text{cm}^1)$ | $\mu_s(1/\text{cm}^1)$ | $\mu'_s(1/\text{cm}^1)$ | $g$ | $n$ | $\delta_{\text{tissue}}(\text{mm})$ |
| --- | --- | --- | --- | --- | --- | --- |
| Skin | 0.18 | 190 | 19 | 0.9 | 1.37 | 130.71 |
| Skull | 0.16 | 160 | 16 | 0.9 | 1.43 | 113.21 |
| CSF | 0.002 | 0.01 | 0.001 | 0.9 | 1.33 | 6.96 |
| Gray Matter | 0.36 | 220 | 22 | 0.9 | 1.37 | 119.26 |
| White Matter | 0.14 | 910 | 91 | 0.9 | 1.37 | 130.11 |

When modeling light diffusion in highly scattering biological tissues, two fundamental assumptions are often made: (1) the absorption coefficient (µ_a_) is considerably smaller than the When modeling light diffusion in highly scattering biological tissues, two fundamental scattering coefficient (µ_s_) and (2) light propagation is isotropic. This isotropic behavior can be expressed using the reduced scattering coefficient µ′_s_ = (1-g) µ_s_ where g is the anisotropic factor^38^. For biological tissues, g typically ranges around 0.9, indicating strong forward-biased scattering ^38^. In terms of light scattering, forward scattering refers to when light is deflected at a small angle, continuing mostly in the same direction as the original beam. This contrasts with backward scattering (light redirected opposite to its initial path) and side scattering (light redirected at a right angle).

#### 2.4.2 Temperature effect: Bioheat Properties and Model Configuration

Modeling of biological thermal systems has been approached on two levels 1) passive section and 2) entire thermoregulatory system, which includes sensors and control mechanisms. The passive section focuses on heat diffusion mechanisms and the associated boundary conditions. Hence, to evaluate the thermal effects caused by absorbed incident light, the Pennes Bioheat Transfer Equation (PBTE) was employed ^26,39,40^. This equation is expressed as follows:

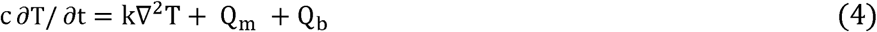

where is the density of the tissue (kg/m^3^), c is specific heat of the tissue (J/kg °C), T is the ambient temperature (25 °C), t is the time (s), k is the thermal conductivity (W/m °C), ∇^2^ is the Laplacian operator, Q_m_ represents heat generation due to metabolic processes within the tissue (e.g., cellular respiration), and is typically assumed to be uniform throughout the tissue (W/m^3^), and Q_b_ represents heat transfer due to blood perfusion, which brings heat into or out of the tissue. (W/m^3^). This is often expressed as proportional to the temperature difference between the tissue and the arterial blood supply:

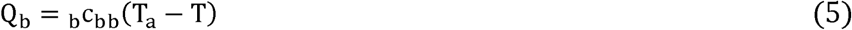

where ρ_b_ is the density of blood (kg/m^3^), c_b_ is the specific heat of blood (J/kg °C), ω_b_ is the blood perfusion rate per unit volume of tissue (1/s), and T_a_ is the arterial blood temperature (37 °C).

Additionally, the heat source term related to absorbed light is defined as:

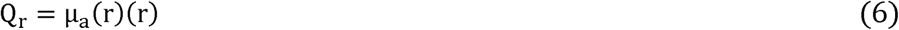

where Q_r_ depends on the local fluence (r) and tissue absorption coefficient µ_a_(r). Equation (5) highlights that heat generation is influenced by the energy available and tissue’s ability to absorb it. Thus, the bioheat transfer equation for continuous wave and pulse wave application are:

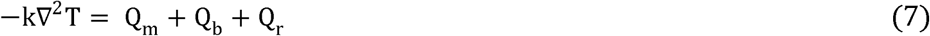

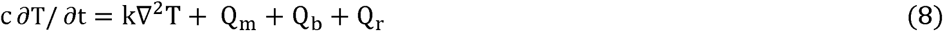

The human thermoregulatory system can maintain a core temperature near 37 °C over a wide range of environmental conditions, even during thermal stress ^34^. Accordingly, the initial tissue and arterial blood temperature were set to 37 °C while subjected to ambient temperature. When the surrounding air temperature is lower than the scalp’s temperature, heat loss (scalp to the environment) occurs to maintain thermal equilibrium via convection. A convective heat flux of 4 W/ m^2^ °C ^41,26^ was used for the stationary surrounding air. Other external boundaries were thermally insulated (i.e. no heat exchange between head tissues and the surroundings).

**Table II** summarizes the thermal properties of tissues, with values adapted from the literature ^42,43,26^.

**TABLE II.** BIOHEAT TRANSFER PROPERTIES.

| Tissue | Thermal conductivity ($\text{W/m } ^\circ\text{C}$) | Specific heat ($\text{J/kg } ^\circ\text{C}^{-1}$) | Density ($\text{kg/m}^3$) | Blood perfusion rate ($1/\text{s}$) | Metabolic heat ($\text{W/m}^3$) |
| --- | --- | --- | --- | --- | --- |
| Skin | 0.37 | 3391 | 1109 | 0.00143 | 363 |
| Skull | 0.32 | 1313 | 1908 | 0.000143 | 70 |
| CSF | 0.57 | 4096 | 1007 | 0 | 0 |
| Gray Matter | 0.55 | 3696 | 1045 | 0.08 | 10437 |
| White Matter | 0.48 | 3583 | 1041 | 0.08 | 10437 |

Noteworthy, PBTE assumes that blood in the capillary bed thermally equilibrates with the host tissue and exits the venous circulation at the local tissue temperature ^44^. This simplification assumes homogeneous blood perfusion, a topic of debate but still a widely accepted standard for predicting heat distribution in biological systems. In fact, recent research by Kabiri et al.^45^, demonstrated that the Pennes equation outperforms the parabolic heat transfer equation in accurately predicting temperature and thermal damage during cranial base neurosurgery.

The American National Standards Institute (ANSI) recommends a surface power density limit of 330 mW/cm^2^ for laser applications ^46^. This limit is predicated based on clinical safe use on the skin ^47,48^. To further relate our predictions to the three power densities of interest, we also determined tissue temperature rises at 330 mW/cm^2^ as a test case.

### 2.5 Finite element computation and Data analysis

The computations were carried out on a workstation equipped with an Intel(R) Core(^TM^) i5-6500 CPU @ 3.20 GHz and 64 GB of RAM. The total computation time was approximately 12 minutes and 27 seconds for continuous wave application (steady-state study) and 48 hours for pulse wave application (transient study).

Our data analysis focused on examining the absorption distribution and temperature profile induced in the brain by tPBM. We characterized these effects using 3-D and 2-D plots, generating graphs that illustrate the absorption and temperature along a straight line from the laser source. Initial estimates of steady-state tissue temperatures were obtained by evaluating the model under prescribed conditions without laser light (i.e., no stimulation), serving as a baseline. We then compared this baseline with the model’s results under laser light at the three power densities (10, 100, and 1000 mW/cm^2^) to assess relative tissue temperature increases. Additionally, we analyzed the power distribution due to 800-nm wavelength.

To elucidate role of tissue optical properties on light transmission, we considered four special cases at 100 mW/cm^2^: (1) uniform properties across all tissues akin to gray matter, (2) all tissues (besides scalp) having equivalent properties akin to gray matter, (3) all tissues (besides scalp and skull) having equivalent properties akin to gray matter, and (4) white matter adopting properties similar to gray matter. Lastly, to investigate the temporal evolution of temperature changes during brain stimulation, we conducted a time-dependent analysis over 20 minutes, maintaining conditions identical to those in the steady-state model.

## 3. Results

### 3.1 Geometry

The geometry considered for the analysis in this study along with laser placement is indicated in **Figure 1**. As mentioned above, the geometry recreates true anatomy reflected by continuous CSF, gyri / sulci specificity, etc.

### 3.2. Steady state analysis: Light propagation

The predicted light propagation is plotted using a combination of 3D surface, 2D cross-section, and line plots in **Figure 2**. Specifically, in **Figure 2A2**, the 3D surface plots reveal a subtle growing “halo-like” effect at the skull surface as light traverses through the scalp. This effect becomes more pronounced at the CSF surface (**Figure 2A3**) that extends to the surfaces of both gray and white matter (**Figure 2A4** and **2A5**). The observed reductions in irradiance as light progresses through the layers are as follows: 1) approximately 65% reduction through the scalp, 2) about 33.9% reduction through the skull, and 3) a minimal reduction of approximately 0.01% through the CSF. Therefore, for a power density of 100 mW/cm², our model predicts that the light intensity reduces by 98.97% at the gray matter surface, yielding an irradiance of 1.03 mW/cm². The corresponding irradiance values for 10 and 1000 mW/cm² reflect a linear trend as expected.

**Figure 2.**
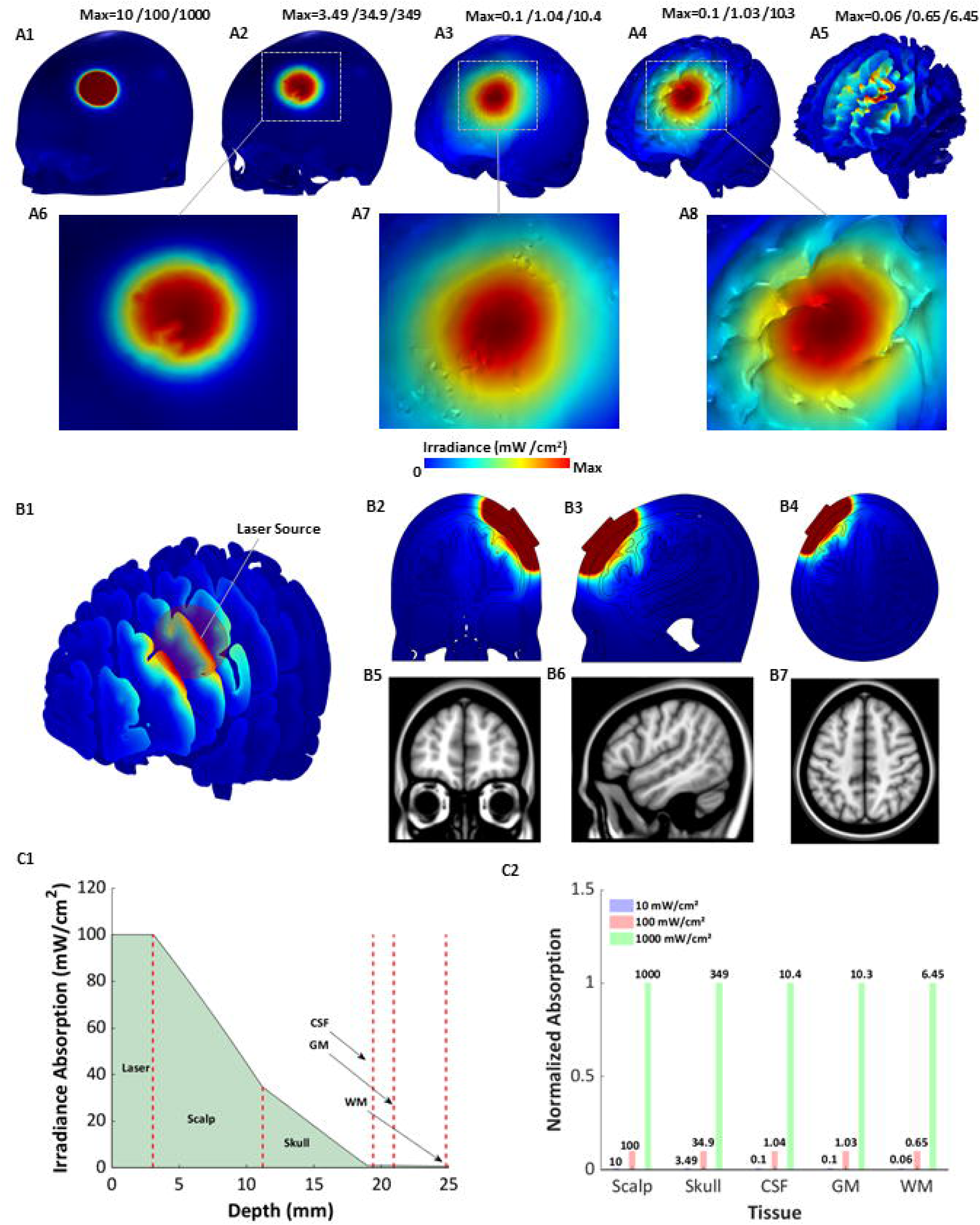
Light propagation from a 800 nm light source positioned at F3 EEG site. *3-D surface* absorption irradiance plots (**A1**:scalp; **A2**: skull; **A3**: CSF; **A4**: gray matter; **A5**: white matter). The predicted maximum irradiance values are noted in the upper right section for each tissue plot. The three values noted correspond to simulation using 10, 100, and 1,000 mW/cm^2^ power density respectively). The dashed sections in **A2, A3, A4** are further expanded in **A6, A7, and A8** highlighting the detail in the model. The growing halo-like effect that is pronounced at the CSF surface is clearly extended to the cortical surface. The irradiance absorption across a series of *2-D cross sectional* slices (**B1**) and individual slices at the center of the laser source highlight depth penetration (**B2-B4**). The images were scaled down to the maximum peak of the gray matter. The corresponding structural MRI slice is also shown (**B5-B7**). Irradiance absorption through an evaluation *cutline* for 100 mW/cm^2^ provides a magnified view of the entire range of the irradiance absorption across head tissues **(C1)**. The evaluation line is shown in Figure 4A inset. The line plot indicates 65% reduction through scalp and 33.9% reduction through skull. The normalized absorption at different power densities is indicated in **C2**. To prevent the dominance of the largest power density over the others, values were scaled down while preserving the relative differences between them.

The expanded sections (**Figure 2A6-2A8)** not only highlight the high resolution detail in the model but help in elucidating the aforementioned halo-like effect.

**Figure 2B2-B4** presents 2D cross-sectional plots illustrating the depth of irradiance absorption. These views (coronal, axial, and sagittal) centered at world coordinates (x = 52, y = 40, z = 48) under the F3 stimulation area reinforce the observations from the 3D surface and the line plot below (**Figure 2C1**). The plot indicates a *rapid* fall-off across the scalp, a *moderate* fall-off across the skull, and a *very gradua*l fall-off at the level of the CSF and brain. For a starting power density of 100 mW/cm², the irradiance reduces to 0.01 mW/cm^2^ (arbitrarily chosen lower value) at ∼134.64 mm from the surface of the scalp and 113.66 mm from the gray matter surface. In **Figure 2C2**, these irradiance values, along with 10 and 1000 mW/cm², have been normalized to aid data visualization. The linear change in values is evident with absorption values in the skull increasing from 3.49 mW/cm² at 10 mW/cm² to 34.9 mW/cm² at 100 mW/cm², reaching 349 mW/cm² at the highest power density.

When discussing irradiance in tPBM, understanding the power distribution across different wavelengths is crucial for optimizing light delivery to the brain. **Table III** reports the power distribution at 800 nm wavelength across the head tissues. Similar to irradiance absorption (see **Fig 2C2**),power distribution follows a linear trendline with varying power density. Considering the surface integral of the applied laser irradiance, the calculated power distribution percentages are as follows: 98.3% (scalp), 30.7% (skull), 3.6% (CSF), 5.7% (gray matter), and 3% (white matter).

**TABLE III.** POWER DISTRIBUTION.

| Tissue | 10 mW/cm<sup>2</sup> | 100 mW/cm<sup>2</sup> | 1000 mW/cm<sup>2</sup> |
| --- | --- | --- | --- |
| Laser | 0.17174 | 1.7174 | 17.174 |
| Scalp | 0.16861 | 1.6881 | 16.861 |
| Skull | 0.052682 | 0.52682 | 5.2682 |
| CSF | 0.0062606 | 0.062606 | 0.62606 |
| Gray Matter | 0.0097759 | 0.097759 | 0.97759 |
| White Matter | 0.0051491 | 0.051491 | 0.51491 |

### 3.3. Steady state analysis: Temperature effects

**Figure 3** depicts the temperature distribution across the three considered power densities, along with their deviations from the condition without stimulation. Across all scenarios, the highest temperatures are concentrated directly beneath the laser source. Moving away from the source, temperatures generally decrease but maintain uniformity throughout most of the brain. At lower power density (10 mW/cm^2^) (**Figure 3B1-B5**), the temperature profile closely resembles the baseline condition. With increasing power density and especially at 1000 mW/cm^2^, distinct increases in temperature emerge. We note maximum temperature increases of 3.76 °C (**Figure 3E1**) at the level of the scalp and 0.57 °C at the gray matter (**Figure 3E4**).

**Figure 3.**
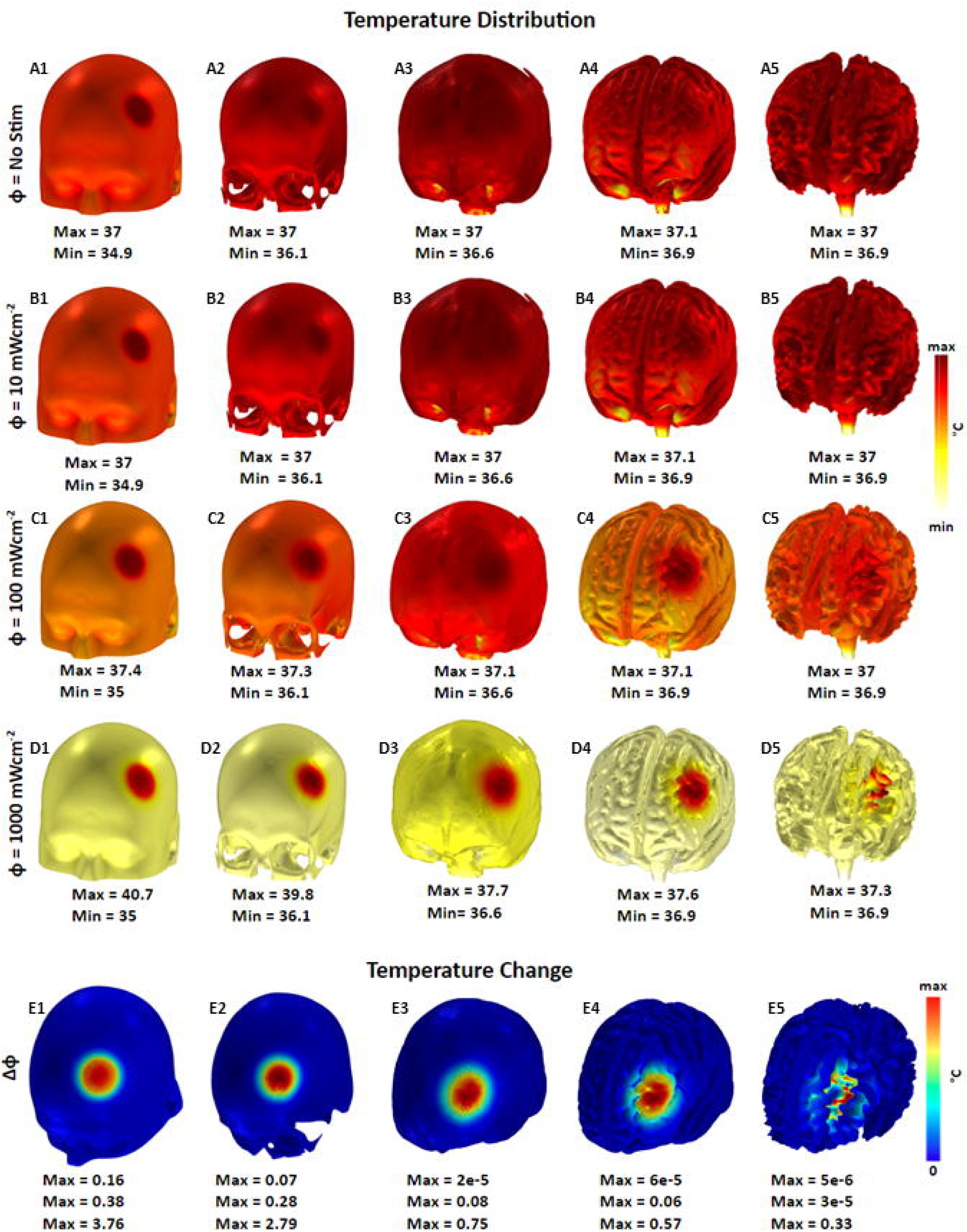
Thermal effects from a 800 nm light source positioned at F3 EEG site. The top row indicates the ‘no stimulation’ condition (**A1-A5)** across scalp, skull, CSF, gray matter, and white matter. The next three rows correspond to light stimulation at 10, 100, and 1,000 mW/cm^2^ respectively (**B1-B5; C1-C5; D1-D5**). The last row denotes the corresponding temperature changes **(E1-E5)**. The predicted temperature rises due to the three power densities are noted immediately below for each tissue plot. A scalp temperature rise of 3.76 °C and brain temperature rise of 0.57 °C is predicted for the 1,000 mW/cm^2^ power density condition.

The temperature variation across each of the head tissues and scalp/brain temperature changes with respect to applied power density is plotted in **Figure 4**. Similar to **Figure 2C1, Figure 4A**,, represents a line plot that crosses normally through all compartments considered in the study at 100 mW/cm^2^. This plot reveals an initial temperature rise followed by a rapid decline across the scalp, a similar fall-off across the skull, dropping slower across CSF, gray matter, white matter and plateauing, thereafter. Additionally, **Figure 4B** shows the minimum and maximum temperatures observed in each tissue at power densities ranging from 10 to 1000 mW/cm^2^. At the lowest power density (10 mW/cm^2^) (blue traces), the maximum temperature stays constant at 37 °C in all tissues, while the minimum temperature at approximately 35 °C at the scalp gradually converges towards 37 °C following a logarithmic growth pattern. Conversely, at higher power densities of (100 and 1000 mW/cm^2^), the maximum temperatures at the scalp are 37.4°C and 40.7°C, respectively. These temperatures drop quickly from the scalp to the CSF region, where they level off and decrease steadily towards 37 °C. The minimum temperature also follows a logarithmic growth pattern in these cases.

**Figure 4.**
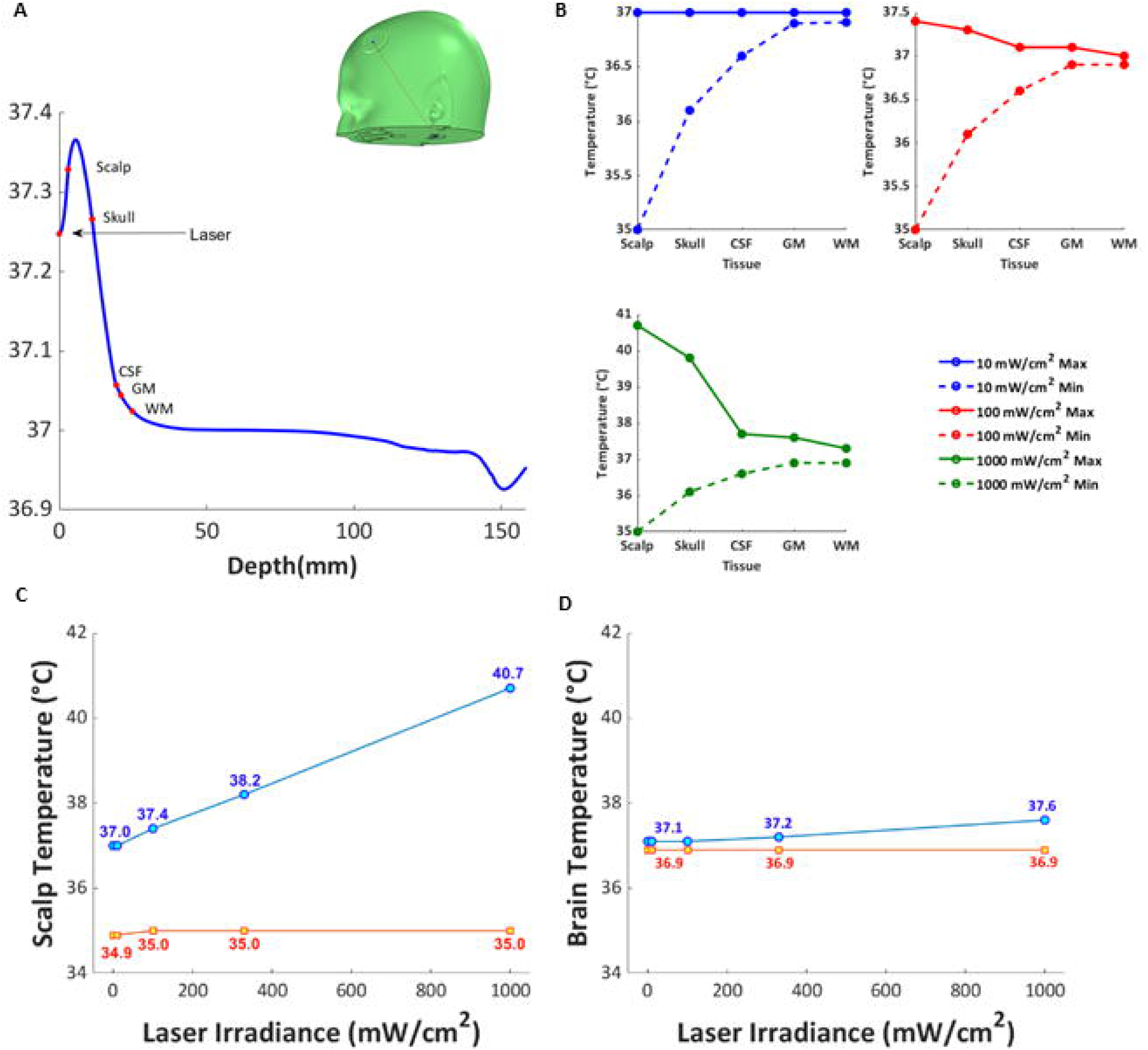
Temperature variation across tissues and across power density. Temperature change variation (maximum) at 100 mW/cm^2^ across the evaluation / cut line **(A**). The cut line extends perpendicularly from the center of the laser source to inside the tissue. See the *inset* in the top right. The maximum and minimum temperature variation at 10, 100, and 1000 mW/cm^2^ **(B)**. Here temperature values correspond to the entire tissue. Predicted scalp temperature as a function of power density **(C)**. Predicted brain temperature as a function of power density **(D**). While increases in power density lead to higher increases in scalp temperature, increases in brain temperature are less rapid. For plots **(C)** and **(D)**, we also include predicted changes at the 330 mW/cm^2^ (ANSI limit) as well.

When plotting scalp temperature with respect to the three applied power densities in addition to the ANSI safety limit (**Figure 4C**), we observe a largely linear relationship with maximum temperature. The corresponding maximum brain temperature rises (**Figure 4D**) are less pronounced, as expected. Finally, minimum temperatures for both scalp and brain are virtually un-changed.

### 3.4 Steady state analysis: Influence of optical properties

The influence of optical properties on spatial and depth profiles of irradiance absorption is plotted in **Figure 5**. The growing halo-like effect at the level of the CSF **(Figure 2A3; 2A7**) is not observed when simulations are performed with homogeneous properties (**Figure 5A1-A6**). When considering original scalp property (**Figure 5B1-B6**), and subsequently, original scalp and skull properties (**Figure 5C1-C6**), the halo-like effect is still not observed. Only when CSF is also assigned its original property (**Figure 5D3**), the growing effect is observed. The effect is further imposed on the underlying layers similar to **Figure 2** above.

**Figure 5.**
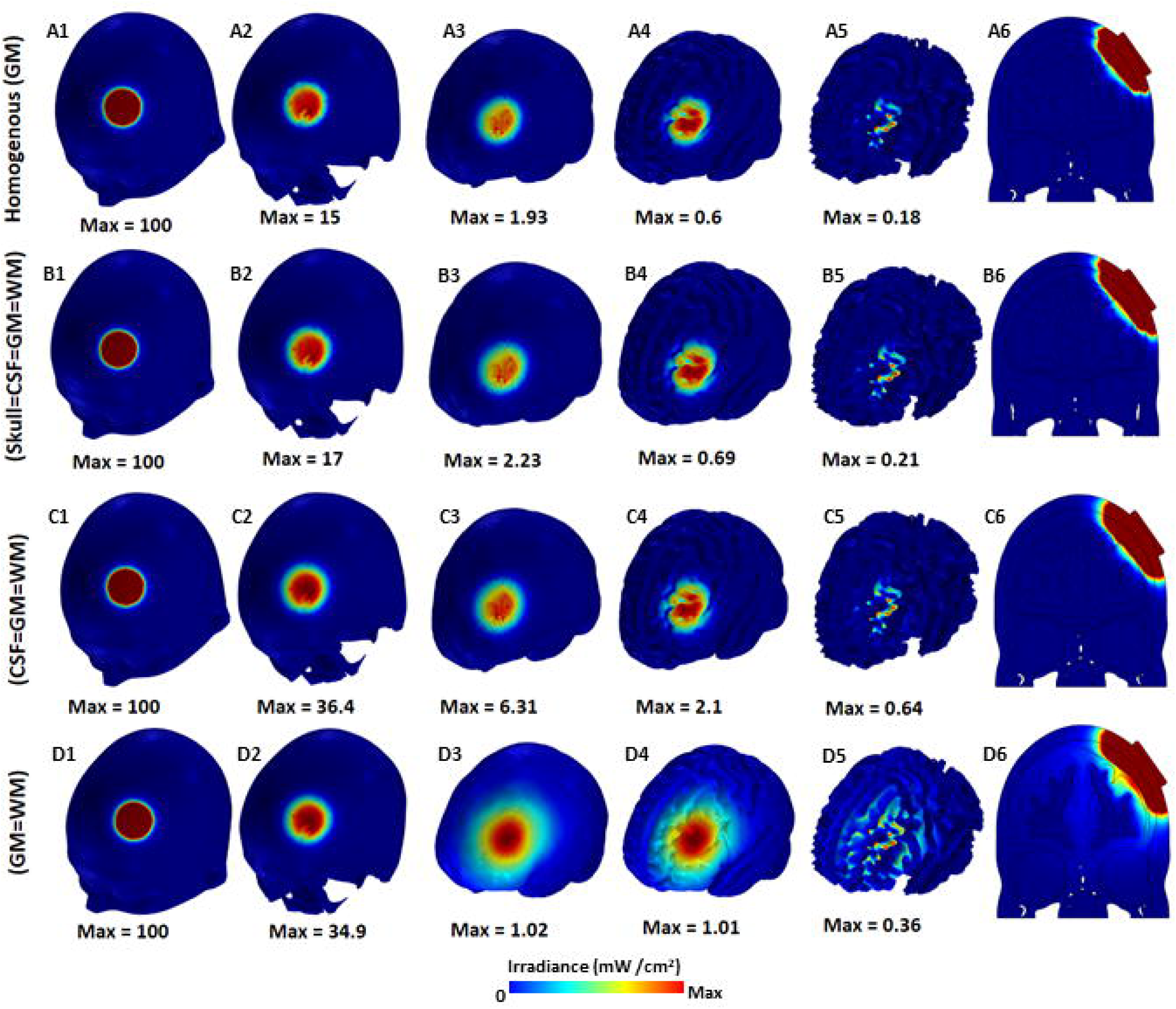
Influence of tissue optical properties on spatial and depth profiles of irradiance absorption. Top row corresponds to simulation results with *homogeneous* optical properties-i.e. properties of all tissues assigned the property of gray matter **(A1-A5**). The second row corresponds to simulation results using the same head model but with all tissues (besides the scalp) having the same optical properties as that of gray matter **(B1-B5**). The third row corresponds to simulation results using the same head model but with CSF and white matter having the same property as that of gray matter **(C1-C5**). The last row corresponds to simulation results using the same head model but with white matter having the same property as that of gray matter **(D1-D5**). This simulation indicates that when the original (true) properties of CSF are re-used, the growing effect is restored. The cross-section profiles corresponding to each of the four aforementioned conditions are plotted in the final column **(A6,B6,C6,D6**). All sectional slices have been scaled down to the max irradiance of the brain.

### 3.5. Transient analysis: Temperature effects

The predicted effects of transient analysis at 100 mW/cm^2^ is plotted in **Figure 6**. Specifically, **Figure 6A** shows the temperature variation as a function of depth within the head for various lengths of time at 100 mW/cm^2^. The model predicts the following temperature increases over a period ranging from 20 to 1200 seconds: ∼0.3 °C (scalp), ∼0.25 °C (skull), ∼0.06 °C (CSF), ∼0.05 °C (gray matter), and ∼0.03 °C (white matter). In **Figure 6B**, which represents the maximum exposure temperature profiles, we note a plateauing effect across all tissues, at approximately 600 seconds (∼10 minutes). Specifically, the scalp temperature reaches a steady state stabilizing at 37.43 °C, while the brain reaches a final temperature of 37.09 °C.

**Figure 6.**
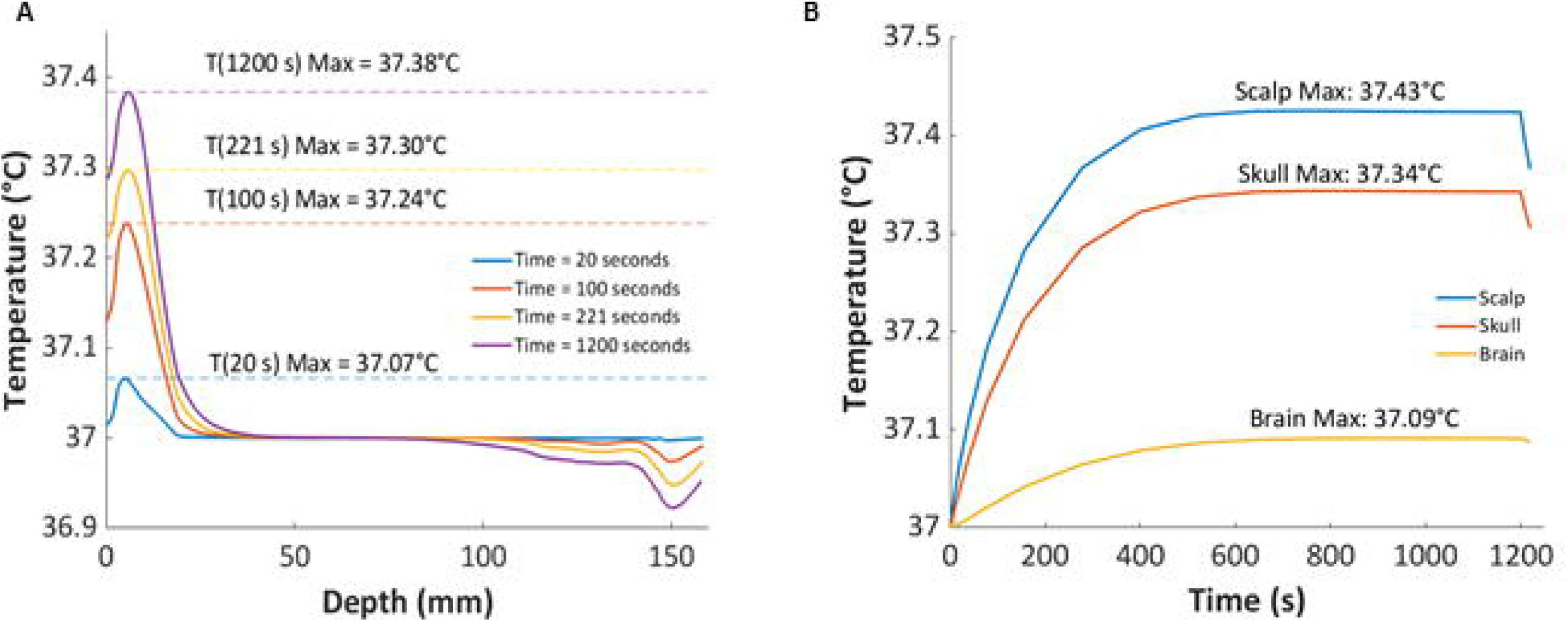
Transient evaluation for 800 nm light with 100 mW/cm^2^ power density. Temperature variation along the cut-line for four different time instants **(A).** As expected, with increasing stimulation runs, scalp temperature increases rapidly. Maximum exposure temperature for the same four time instants **(B)**. The plots indicate that scalp temperature increases plateau after ∼ 9 minutes, whereas the brain temperature increases plateau after ∼ 6 minutes.

The key results pertaining to light penetration and safety considerations (i.e. scalp and brain temperature rises) across the considered power densities are noted in **Table IV**.

**TABLE IV.** RESULTS SUMMARY.

| Power density ($\text{mW/cm}^2$) | Irradiance absorption at Skull ($\text{mW/cm}^2$) | Irradiance absorption at CSF ($\text{mW/cm}^2$) | Irradiance absorption at gray matter ($\text{mW/cm}^2$) | Temperature increase at Scalp ($^\circ\text{C}$) | Temperature increase at Brain ($^\circ\text{C}$) |
| --- | --- | --- | --- | --- | --- |
| 10 | 3.49 | 0.1 | 0.1 | 0.16 | 6e-5 |
| 100 | 34.9 | 1.04 | 1.03 | 0.38 | 0.06 |
| 330*<br>(ANSI<br>limit) | 115 | 3.42 | 3.39 | 1.24 | 0.19 |
| 1000 | 349 | 10.4 | 10.3 | 3.76 | 0.57 |
| 100** | - | - | - | 0.43°C | 0.09 |

\*Indicates data for the ANSI limit. \*\*Indicates transient analysis at 1200 seconds. For these test cases, only temperature increases were determined.

## 4. Discussion

Our study serves as the first transcranial PBM simulation study that considers high resolution 3D geometry (1 mm) with realistic anatomical detail. This is reflected by the geometry considered in the study (**Fig 1)** and subsequently, the enhanced visualization of light propagation through the different tissues (**Fig. 1A6-A8**). Previous efforts have either been limited to reduced geometries (layered slab with slots mimicking the sulcus, layered head model) ^34^ or the ones with 3D geometries involving simplifications (concentric spheres, layered cylinder, fused scalp and skull geometries, missing CSF) ^26,27,33,40^. As mentioned above, our geometry recreates true anatomy-e.g. zygomatic process in skull, eye cavities, continuous CSF, and gyration in gray matter. We further consider true laser source geometry (instead of a point source). The consideration of realistic geometry is a critical step in performing simulations with more accuracy^49^.

Historically, the main sources to demonstrate safety and efficacy of medical devices have been bench, pre-clinical, and clinical studies. With the advent of digital computing, computational modeling has continued to become more powerful, and consequently, being increasingly used to support regulatory decision making^50^. In fact, the US FDA has identified computational modeling as one of the priority areas for the agency moving forward. With respect to tPBM, we strived to address the main questions-the extent of light penetration, induced spatial /depth profile, and associated risk factors (i.e. scalp and brain temperature increases). Our results indicate that for 800 nm wavelength, 1.03% of the surface power density and 5.7% of the applied power is delivered to the gray matter. While the spatial light distribution mimics the shape (circle) and extent (diameter) of the source, there is a noticeable growing or halo-like effect at the level of CSF which is further extended down to the gray matter surface. We verified that the CSF is primarily responsible for this aforementioned effect. This observation was also noted by Okada et al. 1997^51^ when comparing theoretical and experimental readings in the context of NIRS. Notwithstanding, the induced profile remains largely focal or restricted to the same underlying locations corresponding to the source location (i.e. F3). The depth focality plot reveals a shallow profile (**Fig. 2B2, 2B3, 2B4**) demonstrating that tPBM is not generally suited for directly targeting deep structures unless alternate application routes are used ^25^. Further, the gyration of the cortical surface has minimal influence as the circular light distribution continues to be imposed on the surface (**Fig. 2A8)**. This is unlike some other non-invasive brain stimulation modalities where the cortical surface gyration plays an important role in “shaping the delivered dose”. While current hotspots are observed in tES^30^, selective electric field increases are demonstrated in gyral crowns and lips in TMS ^52^.

When evaluating steady-state temperature rises, we noted that a 1,000 mW/cm^2^ surface power density results in a 0.57°C rise in the gray matter. While international regulations based on magnetic resonance and implantable devices indicate cell damage / tissue ablation with focal temperature increases at >2°C, changes in cell excitability are evident starting at 0.5°C ^53,42^. Taken together, our results indicate that the usage of 1,000 mW/cm^2^ power density is not recommended as it risks inducing thermal changes that may affect cellular excitability. Whereas, the usage of 100 mW/cm^2^ and lower should pose no safety concerns, as the predicted temperature rise is an order of magnitude away from the cell property change threshold. The corresponding scalp temperature increase at 1,000 mW/cm^2^ (3.7°C) is also of concern. The US FDA approved Tumor Treating Fields technique for glioblastoma keeps the scalp temperature to 39.5°C and lower to avoid thermal harm ^54^. Therefore, the use of 1,000 mW/cm^2^ is also not recommended based on safe scalp temperature limits. Further, the spatial profile of induced temperature increases mimics that of the light distribution-as one would expect. The same circular shape and restricted to the diameter of the source is clearly observed.

It should be further noted that while the irradiance absorption profiles are linear (i.e. the same increase in surface power density equates to the same increase in brain power density), the induced temperature rises are not. This is expected given the physics of the thermoregulatory mechanism imposed by the Pennes equation. The induced maximum and minimum temperature profiles indicate converging plots as light traverses through different head tissues. At the level of gray matter, while the overall differences are small, they increase with increasing power density (delta= at 10 mW/cm^2^ and = at 1,000 mW/cm^2^).

The transient temperature analysis indicates that the greater the stimulation period, the higher is the induced scalp temperature. The temperature rise in the scalp is rapid and falls equally rapidly as light travels through the tissues. The predicted temperature increase of ∼0.43°C for a period of 1200 seconds (20 min) remains within the safety limit and is similar to the predicted increase in steady state condition (0.38°C). Further, the maximum exposure temperature curves indicate that the temperatures eventually plateau at ∼10 minute mark. This would mean that while increasing stimulation periods would increase the maximum scalp temperature reached in concomitant fashion, any additional increase beyond the 10 minute mark is not predicted. For instance, when employing a 2400 seconds (40 min) protocol, the maximum scalp temperature reached would be higher than 0.43°C but would hold steady beyond the 10 minute mark.

Our results are within the range of results reported in prior 3D computational and validation efforts, in spite of a multitude of differences (geometry, physics, source application, tissue properties). Bhattacharya et al.^26^ notes temperature increase in the brain and scalp of 0.04°C and 0.25°C respectively at 810 nm with a 500 mW/cm^2^ point source positioned at Cz. Ibrahimi et al.^27^, used 810 nm light with 100 mW/cm^2^ source and demonstrated light penetration of 10 mm into the cortex and scalp temperature increase of 0.45°C. Our results indicate reduction of fluence rate to 0.01 mW/cm^2^ at ∼113 mm from the gray matter surface. Yue et al. ^33^ using 850 nm NIR demonstrated penetration depth of ∼120 mm for normalized photon flux of ∼10 e-10 with a single source positioned on the frontal cortex. When comparing experimental efforts, Jagdeo et al. ^18^ study is highly relevant due to the use of cadaver skull with soft tissue and even more so with Morse et al.^21^ study due to the use of a fresh cadaver. The reported 2.1% light penetration at the frontal cortex reported by Jagdeo et al. ^18^ is similar to the 1% of the injected irradiance found in our study. Further, our prediction of irradiance loss at scalp (65%) corroborates with the 50% reported ^21^. When using MR thermometry, while Dmochowski et al.^46^ did not find any significant differences between active and sham arms, they noted a maximum absolute temperature decrease of 0.14°C with active stimulation at 122 seconds post-stimulation. The study used an 808 nm laser source corresponding to 330 mW/cm^2^ directed at the frontal pole for 10 min. While the decrease instead of increase is unexplained, our simulation at 330 mW/cm^2^ predicts a cortical temperature rise of 0.19°C. The authors postulate that the absence of significant differences could indicate that changes were not detectable within the temporal and temperature resolution of measurements. A proton magnetic resonance spectroscopy study further noted no significant brain temperature differences between active and sham tPBM when applying power density of up to 850 mW/cm^2^ at F3/F4 locations^55^. The authors further note the timing of data collection (15 min post tPBM) as a limitation and suggest immediate post-treatment measurement studies. As our simulations indicate that the time course of tissue temperature rises immediately from the illumination onset, a concurrent evaluation would enable a more direct comparison.

Our modeling results can be used to further optimize tPBM stimulation parameters. As the induced light distribution directly mimics the shape and size of the source, one could potentially use a smaller or bigger source to directly impose desired spatial profiles. Further, as the profiles are induced directly under the source, one could potentially use multiple sources to simultaneously target different brain regions. Temperature increases will likely pose no concern as the induced profiles will be co-located with light distribution profiles. Our transient analysis indicates that tissue temperature continues to rise for the first 10 minutes of application. This implies that if a protocol were to employ smaller stimulation times (e.g. intermittent application in on-off fashion), one could potentially avoid the highest possible temperature rise (for a given dose).

On a high level, tPBM demonstrates some similarity to TMS with respect to the generation of a consistent spatial profile dose (i.e. irradiance). In TMS, peak induced EF is induced directly under the intersection of the coil windings, analogous to the peak irradiance under the light source. TMS modeling has however advanced rapidly with precise predictions linked to coil angle and orientation. tPBM is similar to tES, in terms of how the respective dose (fluence and current) needs to account for contribution due to each tissue. However, tES can be optimized to target deeper structures ^56,57^ and has successfully demonstrated this capability^58^. Further, with respect to tFUS^59^, where deep regions can be targeted sparing superficial regions, the benefit of tPBM is its simplicity. That is, the induced cortical light distribution follows the source shape and dimensions.

As with any simulation effort, our predictions are limited by the tissue properties considered in the study. Future efforts should consider a parametric evaluation - i.e. study changes in light diffusion and associated temperature rises due to variation in optical and thermal properties in one anatomical dataset. Similarly, the effect of inter-individual anatomical variation should be explored. However, the imposition of the relatively same irradiance profile (i.e. shape/extent matching source), suggests less impact-to both anatomy and properties. Further, the consideration of an average dataset in our study demonstrates what one would reasonably expect at a population level.

While overall risk analysis of a tPBM system requires evaluation of several other parameters from laser classification, engineering control measures, maximum permissible exposure (MPE), our extensive computational analysis helps address light distribution and associated temperature changes. The consideration of a highly realistic 3D model and related accuracy therefore helps advance the tPBM field. Our predictions are expected to assist not only brain stimulation researchers in planning dosing for their tPBM studies but also safety personnel and readers who are generally interested in tPBM and contemplating pros and cons with respect to other brain stimulation modalities.

## Authorship statement

Mr. Guillen and Drs. Truong and Datta designed the study. Mr. Guillen and Dr. Truong conducted the study, including data collection and data analysis. Mr. Guillen, Mr. de Taboada, and Drs. Truong and Datta performed data interpretation. Mr. Guillen and Dr. Datta prepared the manuscript draft with important intellectual input from Drs. Truong, Faria and Pryor and Mr. de Taboada. Mr. Guillen and Drs. Truong and Datta had complete access to the study data. All authors approved the final manuscript.

## Data Availability

All data produced in the present work are contained in the manuscript

